# Genome-wide analysis and polygenic prediction of clinical obesity and comparison with body mass index

**DOI:** 10.64898/2026.05.20.26353665

**Authors:** Sohail Zahid, Seraj N. Grimes, April Kim, Zhiqi Yao, Allison W. Peng, Roger S. Blumenthal, Rexford S. Ahima, Marios Arvanitis, Michael J. Blaha, Alexis Battle

## Abstract

A new definition of clinical obesity was introduced by the 2025 Lancet Diabetes & Endocrinology Commission to characterize excess body fat and dysfunctional adiposity beyond body mass index (BMI). We performed the first genome-wide analysis (GWAS) of 151,642 cases of clinical obesity and 128,874 controls without obesity from the All of Us Research Program and UK Biobank spanning five ancestry groups and compared these results to GWAS for BMI. We identified 127 independent loci associated with clinical obesity, 63 of which did not share any significant association with BMI. We highlight rs15285 as the most discordant variant, with larger effect size and significance for clinical obesity compared to BMI (ΔZ: +6.35, Δ -log10 P: 17.67). This variant is located in the LPL gene, colocalized to expression quantitative loci for lipoprotein lipase, and associated with elevated triglyceride levels and proteomic markers of insulin resistance and inflammation. Next, we constructed a clinical obesity polygenic score, which had improved association with cardiovascular risk factors and proteomic markers of inflammation (interleukin-6, fibroblast growth factor 21, hepatic growth factor) and insulin resistance (adiponectin, resistin, leptin, and leptin receptor) over a BMI polygenic score. Stratifying individuals into low, intermediate, and high inherited obesity risk groups, we found that clinical obesity polygenic risk reclassified 35% of individuals compared to BMI polygenic risk. Clinical obesity polygenic risk improved discrimination for myocardial infarction, heart failure and stroke over BMI polygenic risk. We replicated the significant improvement in cardiovascular risk prediction of clinical obesity polygenic scores in Atherosclerosis Risk in Communities, Multi-Ethnic Study of Atherosclerosis, Framingham Heart Study, and Women’s Health Initiative. These findings demonstrate that clinical obesity captures genetic loci distinct from BMI that are biologically and clinically relevant to cardiovascular health and can improve cardiovascular genetic risk prediction.

## Manuscript

Obesity is a public health epidemic and a major contributor to morbidity and mortality across the world.^1–3^ While obesity has historically been characterized by body mass index (BMI), this metric can mischaracterize adiposity at the individual level since it does not incorporate body shape, lean mass, physical fitness, or metabolic disease.^4, 5^ To address these limitations, the Lancet Diabetes & Endocrinology Commission introduced a new obesity definition in January 2025 to deprioritize BMI and created a clinical obesity category defined as both excess body fat and adiposity-related dysfunction.^6^ This new definition has spurred significant discussion in the medical community, with recent studies showing clinical obesity is common in the general population, present even with “normal” BMI, and associated with cardiovascular mortality.^7–13^ However, despite these emerging insights, the biological mechanisms underlying clinical obesity and their differences from BMI are not well understood.

In particular, the genetic architecture of clinical obesity is unknown. Prior genome wide association studies (GWAS) of BMI have led to the discovery of hundreds of independently associated variants, most whose function remains unknown.^14, 15^ Broader genomic studies of obesity have understanding by focusing on rare variants, ectopic fat depots on imaging, its uncoupling from cardiometabolic traits, metabolic clusters, or cumulative polygenic risk.^16–22^ By refining diagnosis to identify excess and dysfunctional adiposity, the new clinical obesity definition creates new opportunities for biological discovery.

In this study, we investigate the common genetic variants associated with clinical obesity in participants in the All of Us Research Program and UK Biobank. We (1) identify variants discordantly significant for clinical obesity compared to BMI, (2) derive clinical obesity polygenic scores and evaluate their association with cardiovascular risk factors and metabolism proteomics, and (3) investigate how clinical obesity polygenic risk scores reclassify inherited obesity risk and predict cardiovascular outcomes. To our knowledge, this is the first study on the genetic architecture and polygenic prediction of clinical obesity.

## Results

### GWAS of clinical obesity

We identified participants with clinical obesity and without obesity using the diagnostic criteria from the Lancet Diabetes & Endocrinology Commission in the All of Us Research Program and UK Biobank, as previously described.^6, 9, 10^ There were 151,642 cases with clinical obesity and 128,874 controls without obesity with available genotyping array data after quality control. We partitioned 70% of individuals for GWAS and reserved the remaining 30% for polygenic risk score analysis. Individuals with clinical obesity were, on average, older, more often male, and had greater body size (**Supplementary Table 1**). We categorized individuals across five genetic ancestry groupings: African, Admixed American, East Asian, European, and South Asian (**Supplementary Table 2**).

After genotyping quality control, we tested associations with 16.5 million imputed single nucleotide polymorphisms (SNPs) with a minor allele frequency ≥1%. We analysed each individual cohort using a logistic mixed model with REGENIE^23^ and meta-analysed all studies with a fixed effects inverse-variance model with METAL.^24^ The SNP-based heritability of clinical obesity was h²=0.272 (SE = 0.010), with an LD score regression intercept of 1.086, λ_GC1000_=1.004 and attenuation ratio of 0.11, consistent with a polygenic architecture as opposed to population stratification. Ancestry-stratified heritability estimates were broadly consistent (**Supplementary Table 3**).

In the meta-analysis, there were 127 loci associated with clinical obesity at a p-value threshold of 5×10^−8^ (**Figure 1A, Supplementary Table 4**). Of these loci, 106 were significant exclusively in the European ancestry GWAS, two reached significance in both European and African ancestry, and one was significant across European, African, and Admixed American ancestry. Eighteen loci did not reach genome-wide significance in any ancestry-specific analysis but emerged significant following meta-analysis. Heterogeneity statistics across significant variants revealed generally consistent effect estimates across cohorts, with most loci demonstrating low to moderate heterogeneity (**Supplementary Table 5**).

**Figure 1.**
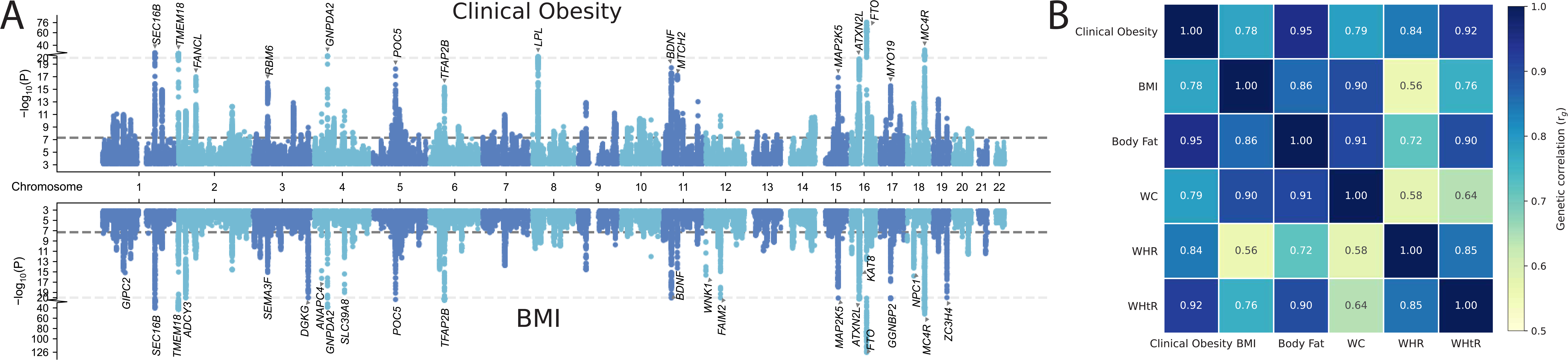
Inherited basis of clinical obesity compared to BMI. (A) Miami plot showing genome-wide significant associations with clinical obesity compared with BMI. Loci meeting a significance threshold of p=10^−15^ are annotated by nearest gene. (B) Genetic correlations of clinical obesity with other anthropometric traits.

The loci with the most significant associations with clinical obesity included variants near FTO (rs56094641, p=7.6×10^−77^), MC4R (rs6567160, p=3.1×10^−33^), SEC16B (rs539515, p=8.0×10^−29^), and TMEM18 (rs12463617, p=2.6×10^−27^), all of which have been reported in prior BMI GWAS. These genes all have known associations with early and severe obesity, involving mechanisms such as hypothalamic appetite regulation, leptin-melanocortinin signalling for energy homeostasis, dietary lipid absorption, and adipogenesis.^18, 25–33^ Tissue and gene set enrichment analyses of genome-wide significant loci for clinical obesity conducted using FUMA highlighted central nervous processes such as those regulating appetite and satiety, consistent with prior genomic analyses on BMI.^14, 15^ The brain was the most enriched organ, with greatest enrichment in the hypothalamus and anterior cingulate cortex (**Supplementary Figure 1**). Cell-type enrichment analysis highlighted the midbrain dopaminergic, GABAergic, and serotonergic populations (**Supplementary Figure 2**). Gene ontology biological process analysis revealed strong enrichment for neuronal differentiation, axon morphogenesis, and synaptic organization (**Supplementary Figure 3**), and canonical pathway analysis showed increased representation of neuronal signalling (**Supplementary Figure 4**).

### Genetic loci discordant between clinical obesity and BMI

To compare the genetic loci for clinical obesity with related anthropometric traits, we conducted multi-ancestry GWAS of body fat percentage, BMI, waist circumference (WC), waist-hip ratio (WHR), and waist-height ratio (WHtR) in the same discovery cohort. All traits showed moderate SNP-based heritability (h²=0.15-0.39) without evidence of population stratification (**Supplementary Table 6**). Clinical obesity was highly genetically correlated with all traits (**Figure 1B**). The largest correlations were with body fat (r_g_=0.95) and WHtR (r_g_=0.92), and the smallest was with BMI (r_g_=0.78).

We subsequently focused on BMI given its widespread role in obesity clinical practice guidelines and public health surveillance, as well as the 2025 Lancet Diabetes & Endocrinology Commission’s call to move beyond this particular metric in favour of clinical obesity.^6, 7^ The clinical obesity and BMI GWAS had similar Manhattan plots with largely overlapping peak signals aside from a few differences (**Figure 1A**). There were 64 loci shared between clinical obesity and BMI, 63 unique to clinical obesity, and 81 unique to BMI (**Supplementary Figure 5**). Among some loci found for clinical obesity but not BMI, previously reported associations have been identified for lipid traits, including triglycerides, high density lipoprotein cholesterol (HDL-C), and low density lipoprotein cholesterol (LDL-C) (**Supplementary Table 7**).

Figure 2A shows a scatter plot comparing loci for clinical obesity and BMI, with top loci more significant for clinical obesity highlighted and annotated by nearest gene. The most discordant variant was located in the 3’ UTR of LPL (rs15285) (Figure 2B). Other loci more significant for clinical obesity include variants near CETP (rs173539), ZPR1 (rs964184), and JMJD1C (rs10740118) (Figure 2C).

**Figure 2.**
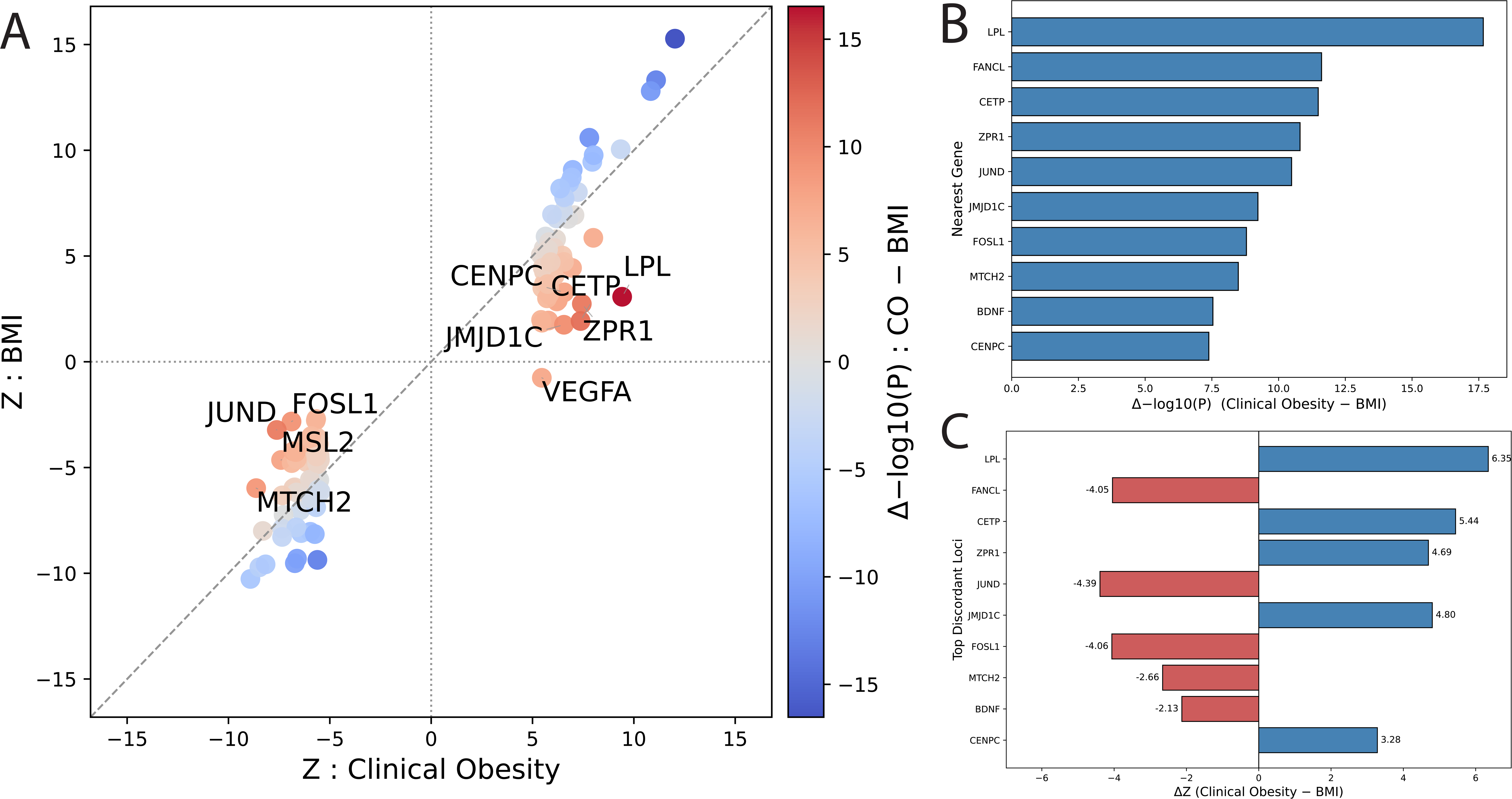
Discordance of clinical obesity loci with BMI. (A) Scatter plot of clinical obesity loci by Z scores for clinical obesity and BMI. (B) Bar chart showing the top loci discordant for clinical obesity compared to BMI by difference in -log(10) p-value. (C) Bar chart showing the difference in Z scores at the top discordant loci. Loci are annotated by nearest gene.

Several of these genes have established mechanistic links to lipid and metabolic function. For example, LPL regulates triglyceride hydrolysis and lipid uptake in adipocyte tissue;^34^ CETP mediates the transfer of cholesteryl esters and triglycerides between lipoproteins;^35^ ZPR1 interacts with high-fat diets, affects neuronal circuits, and influences energy metabolism;^36^ and JMJD1C promotes lipogenesis by demethylating H3K9me2 in response to insulin.^37^ There was one variant near VEGFA (rs9472125) with a sign flip between the clinical obesity and BMI GWAS (Figure 2A), while the clinical obesity effect was concordant with prior significant associations for BMI-adjusted waist circumference and insulin resistance markers.^20, 36, 38^ VEGFA has been shown to regulate adipose tissue vascularization, impacting fibrosis, inflammation, and insulin resistance.^39^

### Lipoprotein lipase on dysfunctional adiposity

We next focused on the LPL locus, which showed the largest discordance in effect sizes and p-values between clinical obesity and BMI (Figure 2C). Regional association analysis at LPL identified robust associations with clinical obesity (Figure 3A). Fine mapping with SuSie identified a single 95% credible set containing 47 variants (**Supplementary Figure 6**). Colocalization analysis with HyPrColoc showed rs15285 colocalized with adipose tissue expression quantitative trait loci from the GTEx v8 data set (posterior inclusion probability= 0.79) (Figure 3B). Heterozygous (C/T) and homozygous (C/C) rs15285 carriers had significantly reduced circulating plasma lipoprotein lipase levels, even after adjusting for BMI or body fat (Figure 3C). In a held-out UK Biobank cohort, rs15285 carriers had significantly increased triglycerides and decreased high density lipoprotein-cholesterol (HDL-C) without significant association with other risk factors (Figure 3D). Among individuals with Olink proteomic data, homozygous rs15285 (C/C) carriers had significantly elevated plasma resistin, interleukin-6 (IL-6), fibroblast growth factor 21 (FGF21), and hepatocyte growth factor (HGF), with leptin receptor trending towards significance (p=0.051) (Figure 3E). These findings suggest that the clinical obesity association at LPL is at least partly mediated by lipoprotein lipase expression in adipose tissue, thereby affecting triglyceride regulation, insulin resistance, and inflammation. This is consistent with the known biology of lipoprotein lipase to hydrolyse triglyceride-rich lipoproteins, generate free fatty acids, and transfer surface remnants to nascent HDL particles.^34, 39^ Accumulation of triglyceride-rich lipoproteins promotes ectopic deposition in liver, activates pro-inflammatory cytokines, and impairs insulin signalling, consequently resulting in metabolic syndrome.^40, 41^

**Figure 3.**
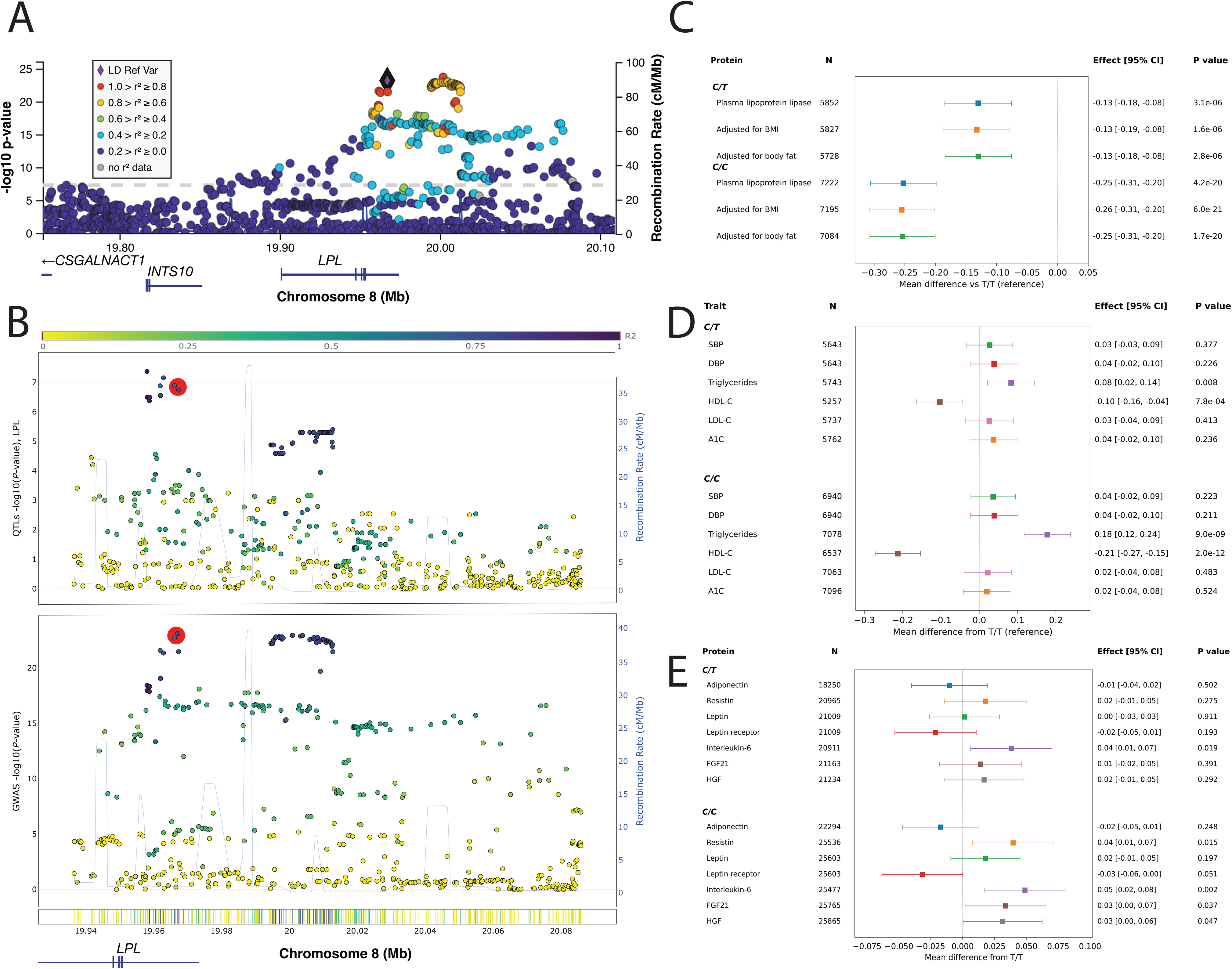
LPL associations with clinical obesity and cardiovascular risk factors. (A) Regional associations at the *LPL* locus with clinical obesity. (B) Colocalization of clinical obesity GWAS with adipose tissue expression quantitative loci for lipoprotein lipase at rs15285. (C) Association of rs15285 carriers with plasma lipoprotein lipase levels. (D) Association of rs15285 carriers with cardiovascular risk factors. (E) Association of rs15285 carriers with proteomic markers of insulin resistance and inflammation.

### Clinical obesity polygenic risk score evaluation

Given the polygenic nature of obesity, we constructed and evaluated a clinical obesity polygenic score (PRS_CO_) using 2.3 million SNPs with LDpred-funct in the held-out UK Biobank cohort. The PRS_CO_ achieved an AUC of 0.65 for discriminating clinical obesity and was correlated with BMI (r=0.268), body fat (r=0.187), and WC (r=0.233) (p < 0.001 for all), explaining 7.2%, 3.5%, and 5.4% of the variance for these traits, respectively. There was a marked gradient in body size across PRS_CO_, with a difference in mean BMI of 6.9 kg/m^2^, body fat of 8.6 %, and WC of 16.7 cm between top and bottom percentiles (Figure 4A**-4C**).

**Figure 4.**
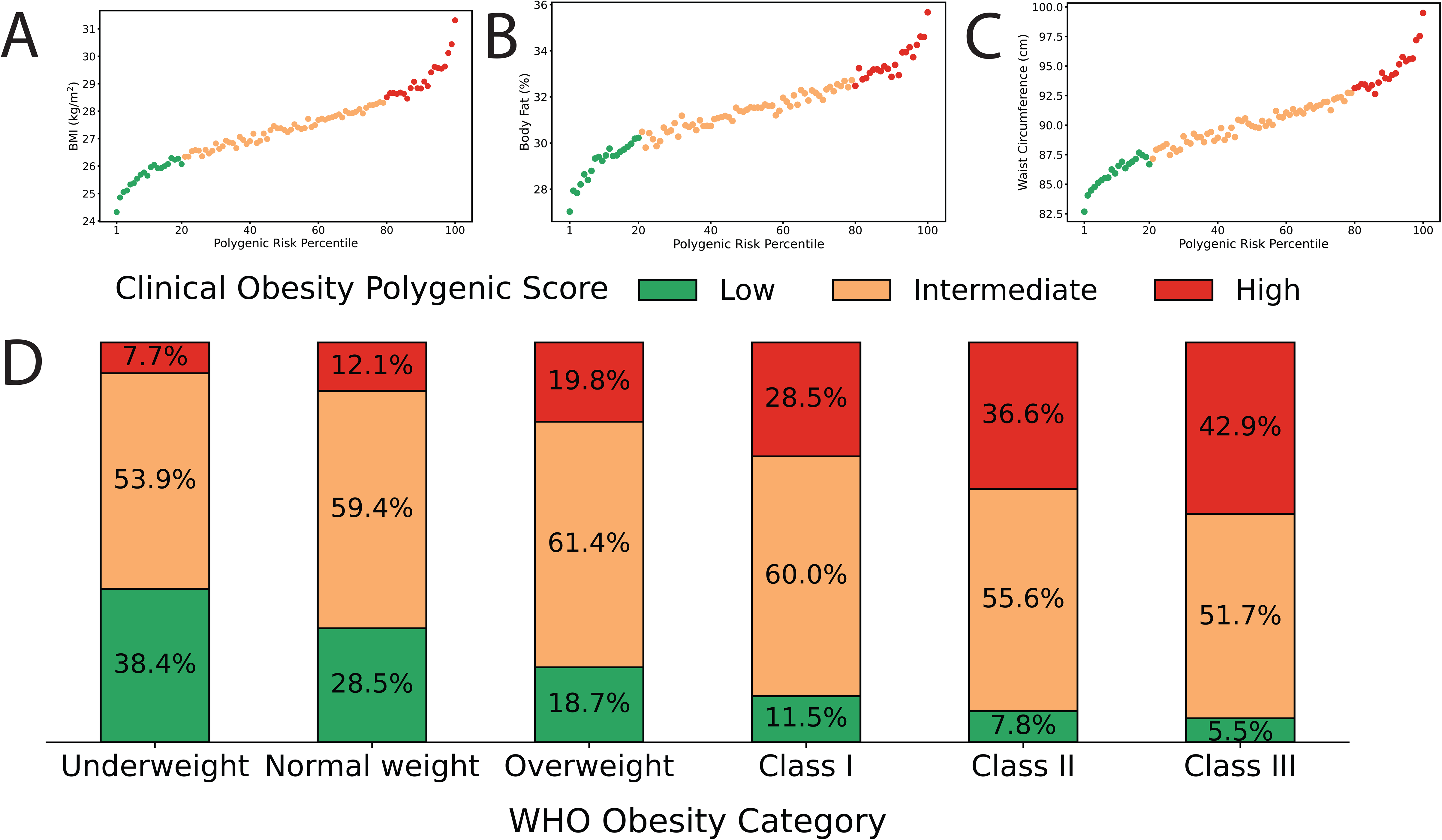
Relationship of clinical obesity polygenic risk with anthropometric traits. Levels of (A) BMI, (B) body fat percentage, and (C) waist circumference across clinical obesity polygenic score percentile, sorted by low (1^st^-20^th^), intermediate (21^st^-80^th^) and high (81^st^-100^th^) groups. (D) Distribution of clinical obesity polygenic risk groups across BMI severity by WHO categories.

We stratified individuals into low (1^st^-20^th^), intermediate (21^st^-80^th^), and high (81^st^-100^th^) PRS_CO_ groups based on percentile. Individuals with high PRS_CO_ had significantly greater BMI, body fat, and WC (**Supplementary Figure 7**). The prevalence of high PRS_CO_ increased progressively across World Health Organization (WHO) BMI categories, reaching 42.9% of individuals with BMI ≥ 40 kg/m² (Figure 4D). However, high PRS_CO_ was not deterministic for obesity, as 19.9% of individuals with high PRS_CO_ had BMI < 30 kg/m^2^ (**Supplementary Figure 8**), underscoring non-genetic factors to body size.

### Cardiovascular risk factor associations

Since the Lancet Diabetes & Endocrinology Commission clinical obesity definition prioritizes identifying dysfunctional adiposity as opposed to excess BMI or body fat alone, we compared PRS_CO_ with polygenic scores for BMI (PRS_BMI_) and body fat (PRS_BF_) generated using LDpred-funct in the same held-out UK Biobank cohort. PRS_CO_ had weaker correlations with BMI (r= 0.268 vs 0.280, p<0.001) and body fat (r=0.187 vs 0.215, p<0.001) compared to PRS_BMI_ and PRS_BFt_ respectively. PRS_CO_ was significantly associated with increased systolic blood pressure (β=0.061, p=1.09×10□□□), diastolic blood pressure (β=0.90, p=5.95×10□¹□¹), haemoglobin A1c (β=0.109, p=7.44×10□²□¹), and triglycerides (β=0.135, p<1×10□³□□), and with decreased HDL cholesterol (β=−0.155, p<1×10□³□□) (Figure 5A). In joint models mutually adjusting for PRS_BMI_ and PRS_BF_, PRS_CO_ demonstrated significant and stronger associations with these cardiovascular risk factors, supporting independent effects beyond BMI and body fat (**Supplementary Table 8, Supplementary Figure 9**).

**Figure 5.**
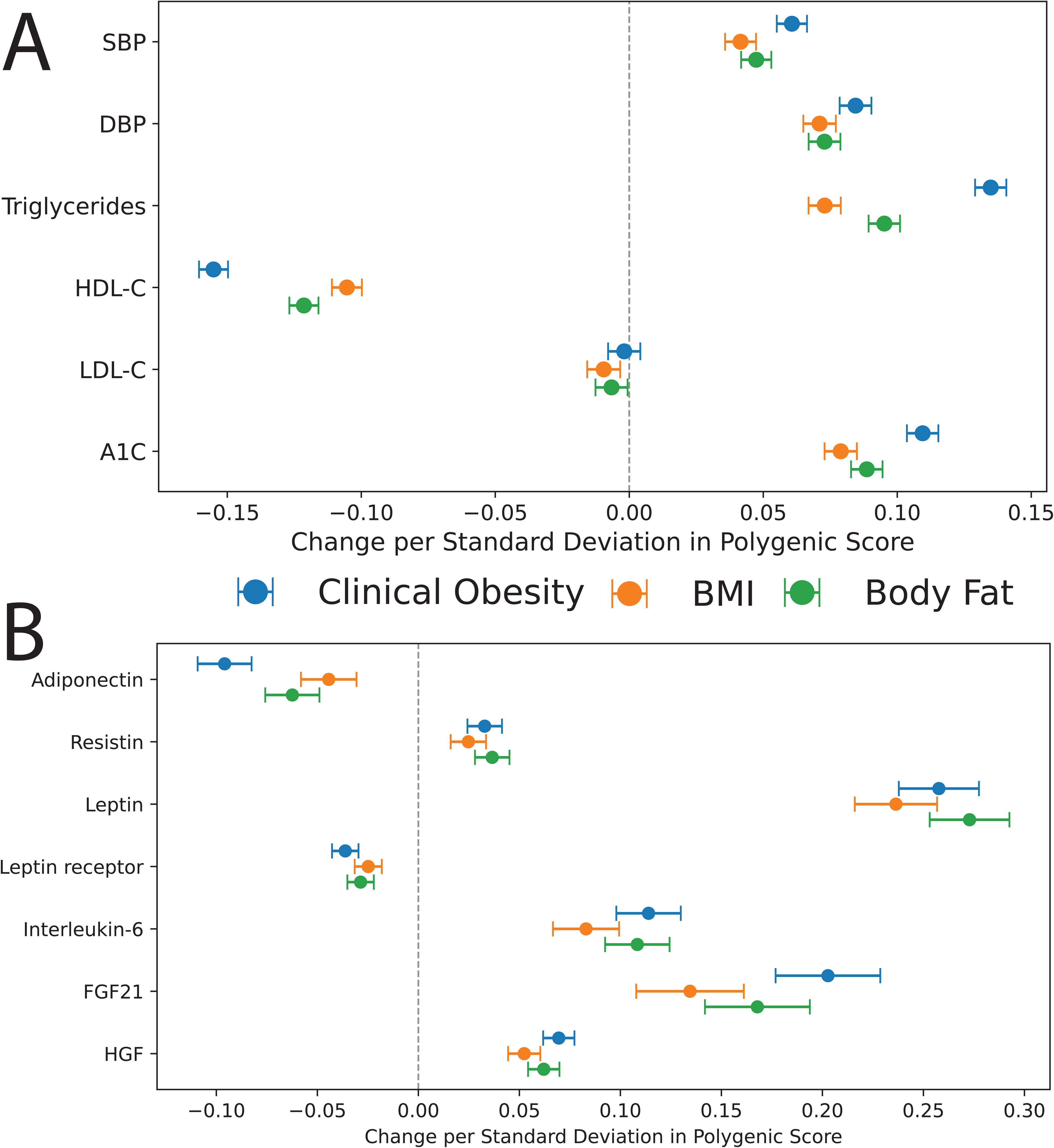
Clinical obesity polygenic risk association with cardiovascular risk factors, insulin resistance, and inflammation. Forest plots showing the association of clinical obesity, BMI, and body fat polygenic scores with (A) cardiovascular risk factors and (B) proteomic markers of insulin resistance and inflammation. Risk factors and proteomic measurements were normalized and assessed per change in standard deviation in polygenic score. Models were adjusted for age, sex, genotyping array, and first 10 principal components of genetic ancestry.

### Proteomic associations with insulin resistance and inflammation

We then evaluated the associations of PRS_CO_, PRS_BMI_, and PRS_BF_ with plasma proteomic markers of insulin resistance and inflammation in the UK Biobank. PRS_CO_ was associated with decreased adiponectin (β=−0.096, P=1.44×10□□□) and leptin receptor (β=−0.036, P=3.20×10□²□), and with increased resistin (β=0.033, P=5.75×10□¹□), leptin (β=0.258, P=3.56×10□¹³□), IL-6 (β=0.114, P=3.76×10□□□), FGF21 (β=0.203, P=1.45×10□□²), and HGF (β=0.070, P=2.35×10□□□) (Figure 5B). After mutual adjustment for PRS_BMI_ and PRS_BF_, PRS_CO_ remained significantly associated with adiponectin, leptin, leptin receptor, IL-6, FGF21, and HGF, with significantly larger effect sizes than PRS_BMI_ and PRS_BF_ for adiponectin, leptin receptor and FGF21 (**Supplementary Table 9, Supplementary Figure 10**).

### Inherited obesity risk reclassification

We next examined how inherited obesity risk classified using different polygenic scores associated with adverse cardiovascular outcomes. Individuals were categorized into low, intermediate, or high risk groups based on PRS_CO_, PRS_BMI_, and PRS_BF_ in the held-out UK Biobank cohort. When comparing PRS_BMI_ with PRS_CO_, most individuals had concordant inherited obesity risk (Figure 6A).

**Figure 6.**
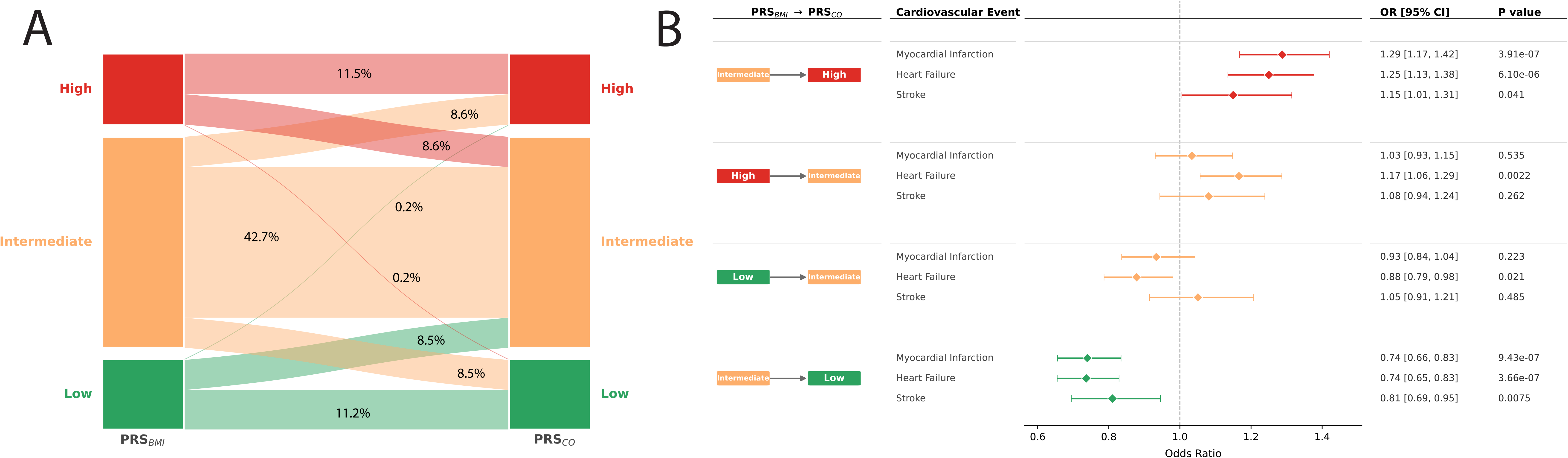
Risk discrimination of clinical obesity polygenic risk with cardiovascular outcomes. (A) Sankey diagram showing the change in inherited obesity risk categorization from BMI to clinical obesity polygenic scores. (B) Forest plots showing the odd ratios of myocardial infarction, heart failure, and stroke when comparing PRS_BMI_ with PRS_CO_ risk groups. Models were adjusted for age, sex, genotyping array, and first 10 principal components of genetic ancestry.

However, 34.6% of individuals were reclassified, primarily between intermediate and adjacent low or high risk categories (Figure 6A). Inherited obesity classification using PRS_CO_ improved cardiovascular risk discrimination for myocardial infarction, heart failure, and stroke compared with PRS_BMI_ (Figures 6B). For example, individuals with intermediate PRS_BMI_ and high PRS_CO_ had increased odds of cardiovascular events whereas those with intermediate PRS_BMI_ and low PRS_CO_ had reduced odds. We similarly compared risk reclassification between PRS_BMI_ and PRS_BF_ and between PRS_BF_ and PRS_CO_. A greater proportion of individuals were reclassified between PRS_BMI_ and PRS_BF_ (42.7%) than between PRS_BF_ and PRS_CO_ (33.7%) (**Supplementary Figure 11**). There also was improvement in cardiovascular risk discrimination from PRS_BMI_ to PRS_BF_ and from PRS_BF_ to PRS_CO_, but these differences were less pronounced than PRS_BMI_ and PRS_CO_ (**Supplementary Figure 12**).

### External validation of improved risk prediction in prospective cohorts

We externally validated our findings in four prospective cohorts with adjudicated cardiovascular outcomes comprising individuals with available genotype data in Atherosclerosis Risk in Communities (ARIC, n=6,950), Multi-Ethnic Study of Atherosclerosis (MESA, n=2,450), Framingham Heart Study (FHS, n=3,179), and Women’s Health Initiative (WHI, n=4,140). In meta-analyses of these cohorts, PRS_CO_ was similarly associated with cardiovascular risk factors: increased systolic blood pressure (β=0.082, p=1.48×10^−25^), diastolic blood pressure (β=0.049, p=1.92×10□^9^), triglycerides (β=0.121, p=8.72×10^−38^), LDL-C (β=0.032, p=1.20×10□³), and fasting glucose (β=0.108, p=4.63×10□^28^), and decreased HDL-C (β=−0.124, p=2.52×10□^49^) (Figure 7A). After mutual adjustment for PRS_BMI_ and PRS_BF_, PRS_CO_ maintained significant association with these cardiovascular risk factors (**Supplementary Table 10, Supplementary Figure 13**). PRS_CO_ had significantly stronger effect sizes with triglycerides and HDL-C compared to PRS_BMI_, suggesting that impaired triglyceride metabolism is more prominent in genetic susceptibility to clinical obesity than elevated BMI.

**Figure 7.**
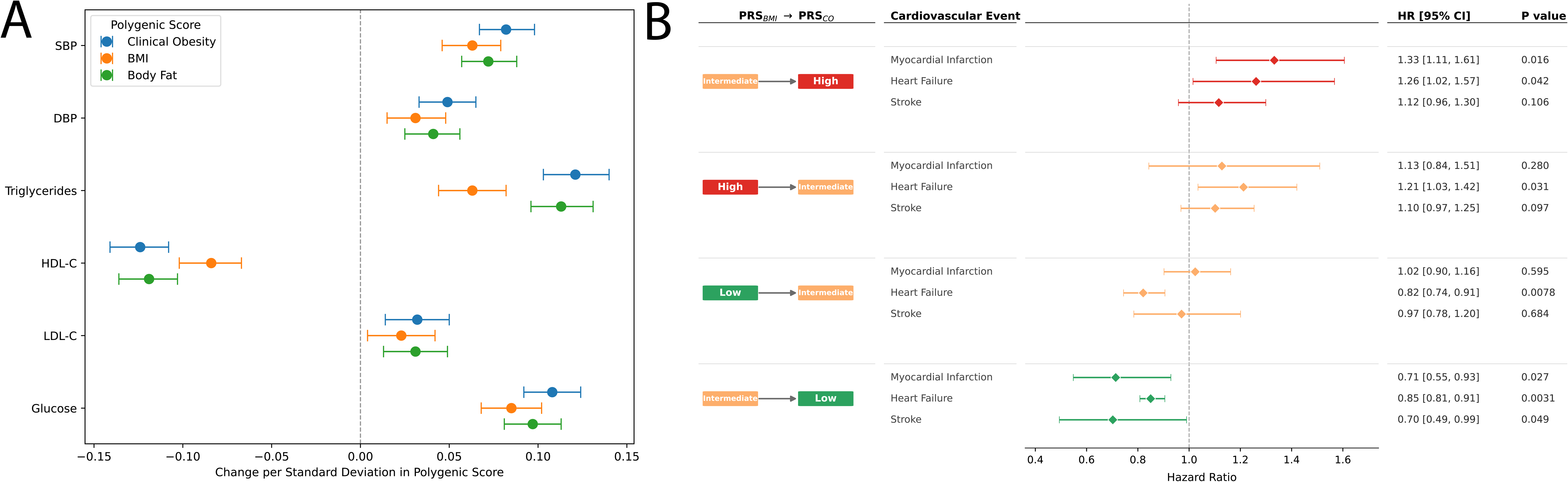
External validation of clinical obesity polygenic risk assessment. (A) Forest plots showing the association of clinical obesity, BMI, and body fat polygenic scores with cardiovascular risk factors in a meta-analysis comprising ARIC, MESA, FHS, and WHI. (B) Forest plots showing the hazard of myocardial infarction, heart failure, and stroke when comparing PRS_BMI_ with PRS_CO_ risk groups. Models were adjusted for age, sex, genotyping array, and first 10 principal components of genetic ancestry.

In prospective meta-analyses across these cohorts, PRS_CO_ was associated with a significantly greater hazard of myocardial infarction (HR 1.24, 95% CI 1.18–1.30; p=1.23×10□¹□), heart failure (HR 1.27, 95% CI 1.21–1.33; p=1.59×10□²□), and stroke (HR 1.17, 95% CI 1.10–1.25; p=3.06×10□□) (**Supplementary Table 11**). In each cohort, we classified individuals into low, intermediate, and high inherited obesity risk based on PRS_CO_ and PRS_BMI_. Risk stratification with PRS_CO_ improved discrimination for elevated hazard for myocardial infarction, heart failure, and stroke (Figure 7B). The magnitude and direction of associations for cardiovascular risk factors and outcomes were consistent across individual cohorts (**Supplementary Tables 10-11**). Together, these findings indicate that inherited susceptibility to clinical obesity captures cardiovascular risk better than genetic BMI risk alone.

## Discussion

In this study, we performed the first genome-wide association study of clinical obesity, identifying 127 significant loci in a large, multi-ancestry cohort of 151,642 cases and 128,874 controls. Among these, 63 loci were specific to clinical obesity and not BMI, including the most discordant variant lying in the LPL locus, highlighting distinct genetic determinants of adiposity-related dysfunction. Finally, we constructed a clinical obesity polygenic risk score using 2.3 million variants that showed stronger associations with cardiovascular risk factors and outcomes across prospective cohorts. These results provide important insights into the inherited basis for clinical obesity and its distinction from BMI.

Our findings reinforce the importance of central nervous system processes in clinical obesity. The top clinical obesity loci (FTO, MC4R, and TMEM18) are widely reproduced in prior genetics studies and have established roles in appetite regulation through the leptin-melanocortin pathway.^14, 15^ Enrichment analyses further highlighted neuronal circuitry particularly in the hypothalamus, anterior cingulate cortex, and midbrain, which collective regulate hormonal satiety signals, cognitive control of eating, and reward processing.^42–44^ These regions are also significantly affected by current obesity pharmacotherapies such as GLP-1 agonists, which act on mesolimbic circuits to attenuate food intake and reduce appetite.^45^

We next found that the clinical obesity loci emphasize peripheral pathways related to triglyceride handling. For example, the top discordant loci between clinical obesity and BMI included LPL, CETP, ZPR1, and JMJD1C, which directly impact triglyceride lipogenesis, homeostasis, and breakdown.^41, 46^ Moreover, PRS_CO_ significantly associates with triglyceride and HDL-C levels more than PRS_BMI_ in multiple, independent prospective cohorts, emphasizing the contribution of triglyceride-rich lipoproteins in clinical obesity genetics. Obesity affects triglycerides through dysregulation of production, lipolysis, remodelling, and clearance.^41, 46–48^ Persistent triglyceride elevations lead to atheroma formation, independently increasing risk for myocardial infarction and ischemic stroke.^40, 49^

The most discordant locus between clinical obesity and BMI was an LPL variant (rs15285), which has been shown to affect LPL mRNA^50, 51^ and was associated with significant differences in lipoprotein lipase, triglyceride, insulin resistance, and inflammation levels in our study. Lipoprotein lipase is significantly elevated in obesity and plays a central role in lipid metabolism by hydrolysing triglycerides in circulating lipoproteins and delivering fatty acids to adipose tissue and skeletal muscle.^34, 52–54^ However, loss-of-function LPL variants contribute to impaired triglyceride clearance, ectopic fat deposition, and increased diabetes and coronary artery disease.^55^ Novel therapeutics targeting lipoprotein lipase via inhibitors of apoC-III (olezarsen, plozarison) and ANGPTL3 (zodasiran, vinacumab) demonstrate significant reductions in triglycerides, hepatic fat, and pancreatitis.^56^ Ongoing cardiovascular outcomes trials will determine whether these agents reduce atherosclerotic cardiovascular events.

We constructed a clinical obesity polygenic risk score (PRS_CO_), extending prior work showing that common genetic variants can collectively confer a significant burden to obesity and related complications.^21^ Individuals with elevated PRS_CO_ had significantly increased body size, metabolic risk factors, insulin resistance, inflammation, and adverse cardiovascular outcomes. These results reiterate that obesity is an important contributor to cardiovascular disease and underscore the value of inherited clinical obesity risk assessment for prevention.^2, 57^ This is especially important as PRS_CO_ was not deterministic for obesity severity, reinforcing studies that lifestyle behaviours can offset elevated genetic risk.^58, 59^

Finally, we showed that PRS_CO_ improved risk discrimination for cardiovascular outcomes, with even greater improvement over PRS_BMI_ than PRS_BF_. These results are consistent with recent genetic evidence that variants increasing BMI do not necessarily confer equivalent cardiovascular risk.^20, 60^ Common genetic variants that uncouple adiposity from its cardiometabolic comorbidities have been shown to implicate peripheral pathways such as lipid metabolism, inflammation, and insulin signalling.^20^ Moreover, monogenic obesity variants with favourable triglyceride handling reduce cardiovascular disease.^60^ These findings support why comprehensive metabolic risk scores outperform BMI in predicting obesity multimorbidity.^19^

Nonetheless, the clinical utility of inherited obesity polygenic scores remain under active investigation, as the clinical obesity definition itself is undergoing critical appraisal for contemporary practice.^61, 62^ Unlike rare variant panel testing which already guides targeted therapy in monogenic obesity, further work is needed to determine the incremental benefit of clinical obesity polygenic scores for diagnosis, risk stratification, or management beyond BMI-based assessment.

Our study has several limitations that must be acknowledged. While we incorporated multi-ancestry data for our genomic analyses, most individuals in our study were still from European ancestry, which reduces generalizability to other populations. Additionally, electronic health records were used to classify clinical obesity diagnoses, which has the potential to misclassify individuals, especially those with underreported or stigmatized conditions.^63, 64^ These limitations are in part a reflection of a broader societal problem with obesity care, in which individuals with obesity face weight-related stigma in clinical settings, delayed diagnoses, and significant healthcare disparities across ethnic and socioeconomic lines.

In conclusion, we carried out genetic association analyses and polygenic prediction of clinical obesity. These results lay the scientific foundation for future variant-to-function studies and investigations into the biological pathways that govern clinical obesity adiposity dysfunction.

## Methods

### Study population

The participants in this study were from two large observational population-based cohorts: the UK Biobank and the All of Us Research Program. The UK Biobank is a prospective study comprising over 500,000 individuals aged 40-69 across 21 assessment centres in the United Kingdom enrolled between 2006 to 2010.^65^ The All of Us Research Program is a prospective study comprising over 400,000 individuals aged 18 and older across more than 340 sites in the United States, with a deliberate focus on recruiting groups traditionally underrepresented in research.^66^ Demographic information, questionnaires, anthropic measurements, laboratory data, imaging, and electronic health records were collected for each participant, as previously described.^65, 66^

### Clinical obesity derivation

We included 151,642 cases with clinical obesity and 128,874 controls without obesity in the UK Biobank and All of Us Research program, as detailed and validated in prior publications.^9, 10^ Clinical obesity was defined using criteria from the Lancet Diabetes & Endocrinology Commission. Briefly, obesity was defined based on body fat percentage from direct measurement, at least two different anthropic measures of excess body size, or severe BMI elevation at least 40 kg/m^2^. Individuals were further stratified as having clinical obesity dependent on the presence of physical impairment or obesity-related dysfunction based on ICD-10 code diagnoses, SNOMED-CT diagnoses, laboratory values, and clinical measurements.

### Genotyping, imputation, and quality control

Genotyping in the UK Biobank was performed using the UK BiLEVE and UK Biobank Axiom arrays and in All of Us Research Program using the Illumina Global Diversity array. Imputation for each cohort was conducted using the Trans-Omics for Precision Medicine (TOPMed) reference panel. Post-imputation filtering was applied to retain variants with an imputation quality score (Rsq) ≥ 0.3 and minor allele frequency ≥ 1%. Quality control procedures were applied consistently across cohorts and included testing for batch, plate, and array effects, deviation from Hardy-Weinberg equilibrium, and discordance across control replicates. Samples were removed for poor genotyping quality, elevated missingness, outlying heterozygosity, relatedness, or sex discordance between genetically inferred and reported sex.

Genetic ancestry was inferred for each participant by principal component analyses as previously described, and assigned to one of five ancestry groups: African, Admixed American, East Asian, European, and South Asian. For individuals in the UK Biobank, we randomly partitioned the dataset into a 70% GWAS discovery set and a 30% held-out set for polygenic score derivation and testing.

### Common variant association studies

Within-cohort GWAS for clinical obesity and anthropometric traits were performed separately for each ancestry group using REGENIE (v4.1), adjusting for age, sex, genotyping array and first 15 principal components of genetic ancestry. Clinical obesity was analysed as a binary trait using a logistic mixed model. Continuous anthropometric traits (body fat percentage, WC, WHR, WHtR) were analysed using a linear mixed model on rank-based inverse normalized values. Step 1 of REGENIE used high-quality genotyped variants (MAF ≥ 1%, MAC ≥ 100, genotyping rate ≥ 99%, HWE p ≥ 10^−15^) after LD pruning (r^2^ < 0.9, 1,000-kb window). Step 2 tested imputed variants with INFO ≥ 0.3 and MAF ≥ 0.001. Summary statistics were combined across cohorts and ancestry groups with fixed-effects-inverse-variance-weighted meta-analyses in METAL. Genome-wide significance was defined as p < 5 × 10^−8^. Independent lead variants were identified by LD clumping (r^2^ < 0.01, 1-Mb window). SNP-based heritability and cross-trait genetic correlations were estimated using LD score regression (LDSC v1.0.1). Fine-mapping was performed using SuSiE to generate 95% credible sets. Colocalization between GWAS signals and adipose tissue GTEx v8 eQTLs were assessed using HyPrColoc.

### Tissue enrichment and pathway analyses

Tissue and gene-set enrichment were performed using MAGMA (v1.10) within FUMA (v1.6.1). Lead variants were annotated to the nearest gene. Tissue enrichment was assessed using MAGMA gene-property analysis against GTEX v8 expression across 54 tissue types. Gene-set enrichment was assessed using the MAGMA competitive model against curated gene sets from Gene Ontology Biological Process, Reactome, KEGG, and WikiPathways. All analyses used a Benjamini-Hochberg FDR threshold of 0.05.

### Polygenic score construction

Polygenic scores were derived using LDpred-funct, which incorporates functional priors to estimate posterior mean SNP effect size for each variant. Linkage disequilibrium was estimated using unrelated individuals of European ancestry from 1000 Genomes Phase 3 as the LD reference panel. The PRS_CO_, PRS_BMI_, and PRS_BF_ were derived in individuals of European ancestry in the held-out UK Biobank cohort using discovery meta-analyses summary statistics for each respective trait and normalized to mean 0 and standard deviation 1.

### Statistical analyses

Associations of rs15285 with cardiometabolic traits and plasma protein levels were evaluated using linear regression in UK Biobank participants of European ancestry. Heterozygous (C/T) and homozygous alternate (C/C) genotypes were each compared to the homozygous reference (T/T) using indicator variables. Cardiometabolic traits included systolic and diastolic blood pressure, haemoglobin A1c, blood glucose, triglycerides, HDL-C, and LDL-C. Proteins included lipoprotein lipase, adiponectin, resistin, leptin, leptin receptor, interleukin-6, FGF-21, GDF-15, and HGF measured in the Olink Explore 3072 platform. All continuous outcomes were rank-based inverse-normal transformed prior to analysis, and models were adjusted for age, sex, batch, and first 10 principal components of genetic ancestry.

Associations of PRS_CO_, PRS_BMI_, and PRS_BF_ with cardiometabolic traits and plasma protein levels were evaluated using linear regression in the held-out UK Biobank and proteomic cohorts respectively. Effect sizes are reported per one standard deviation increase in polygenic score. For risk stratification analyses, participants were ranked by each polygenic score and categorized into low (1^st^ - 20^th^ percentile), intermediate (21^st −^ 80^th^ percentile), and high (81^st −^ 100^th^ percentile) risk groups. To compare inherited risk across adiposity measures, participants were jointly classified using pairwise combination of scores: PRS_BMI_ & PRS_CO,_ PRS_BF_ & PRS_CO,_ and PRS_BMI_ & PRS_BF_. Myocardial infarction, heart failure and stroke were ascertained using algorithmically-defined phenotypes and ICD-10 diagnosis codes. Logistic regression was used to estimate the odds of each cardiovascular outcome across risk categories referenced against the joint intermediate group, adjusted for age, sex, genotyping array, and the first 10 principal components of genetic ancestry.

## External validation

To assess generalizability of polygenic score performance, we performed external validation in four independent, external prospective cohort studies with adjudicated cardiovascular outcomes from Atherosclerosis Risk in Communities (ARIC), Multi-Ethnic Study of Atherosclerosis (MESA), Framingham Heart Study (FHS), and Women’s Health Initiative (WHI). We generated PRS_CO_, PRS_BMI_, and PRS_BF_ in unrelated individuals of European ancestry for each cohort using LDpred-funct as described above. Associations of each polygenic score with fasting blood glucose, systolic and diastolic blood pressure, triglycerides, HDL-C, and LDL-C were evaluated by linear regression. Associations with incident myocardial infarction, heart failure, and stroke were evaluated using Cox proportional hazard regressions, with follow-up time defined from cohort enrolment to first outcome occurrence, death or end of follow-up, whichever occurred first. All associations were reported per one standard deviation increase in polygenic score and adjusted for age, sex, and first 10 principal components of genetic ancestry. For risk stratification analyses, participants were categorized into low (1^st^ - 20^th^ percentile), intermediate (21^st −^ 80^th^ percentile), and high (81^st −^ 100^th^ percentile) PRS_CO_ and PRS_BMI_ risk groups and then jointly classified into pairwise risk categories. The hazard of each cardiovascular outcome was estimated across joint risk categories relative to the joint intermediate group using Cox proportional hazards regression. Cohort-specific estimates were pooled by fixed-effects inverse-variance-weighted meta-analysis

## Ethical approval

UK Biobank data was approved though application number 98985 in the UK Biobank. All of Us Research Program data was approved though application rw-9d5cde2b. Data from Atherosclerosis Risk in Communities Study, Multi-Ethnic Study of Atherosclerosis, Women’s Health Initiative, and Framingham Heart Study were downloaded from dbGAP (accession numbers: phs000007.v29.p11, phs000287.v6.p1, phs000209.v13.p3, phs000280.v4.p1, phs000200.v11.p3, phs000888.v1.p1). All individuals provided informed consent for participation in the individual cohorts. Our GWAS study was approved by the Johns Hopkins School of Medicine IRB.

## Data Availability

UK Biobank data are accessible through the application process outlined at https://www.ukbiobank.ac.uk/enable-your-research. Detailed information on the genetic data provided by UK Biobank is available at http://www.ukbiobank.ac.uk/scientists-3/genetic-data/ and http://biobank.ctsu.ox.ac.uk/crystal/label.cgi?id=100314. All of Us Research Program data are accessible to registered researchers through the Researcher Workbench at https://www.researchallofus.org. Data from ARIC, MESA, FHS, and WHI are available through controlled-access application via the NIH database of Genotypes and Phenotypes (dbGaP) at https://www.ncbi.nlm.nih.gov/gap.

