## Supplementary figures and images for "Genome-wide analysis and polygenic prediction of clinical obesity and comparison with body mass index"

### Figure S1

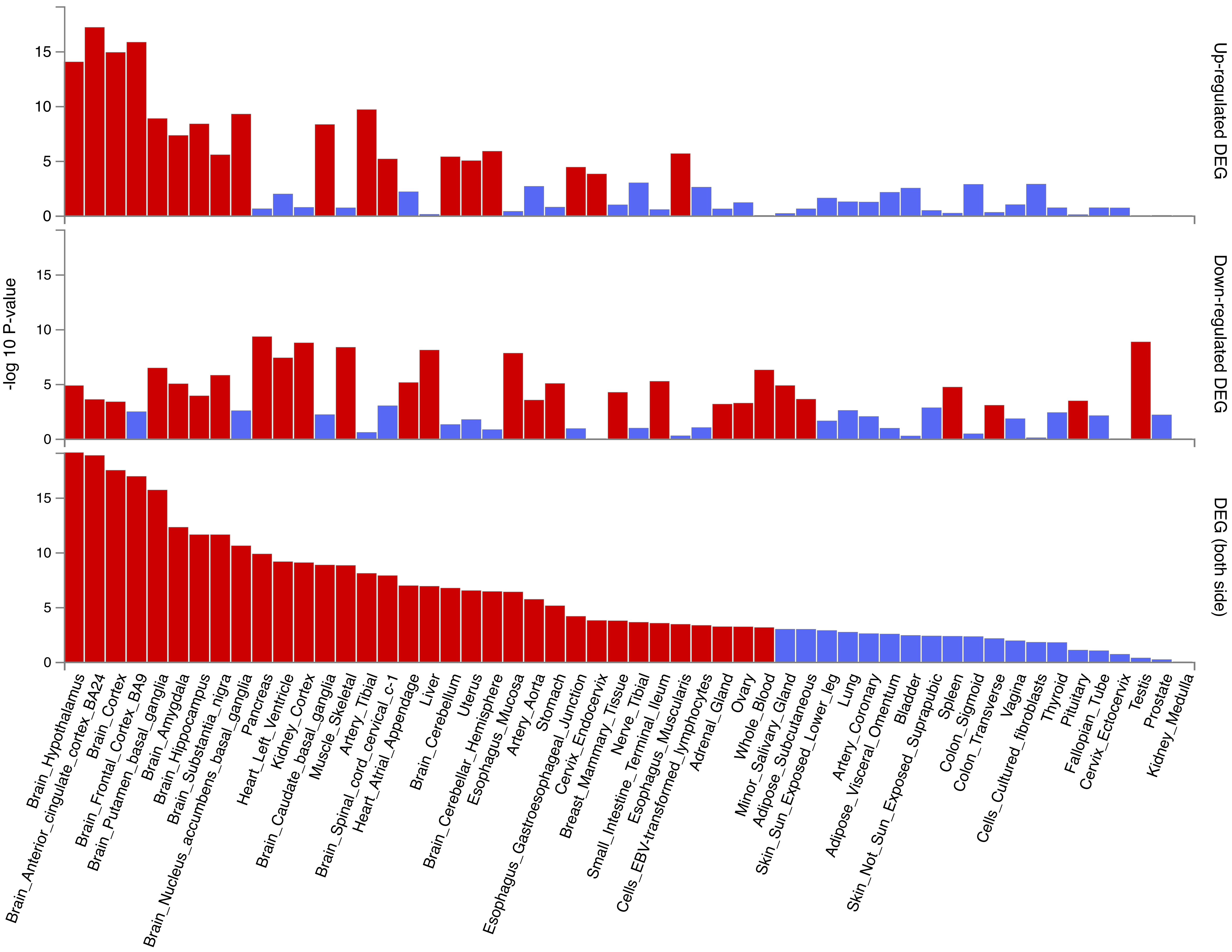

### Figure S2

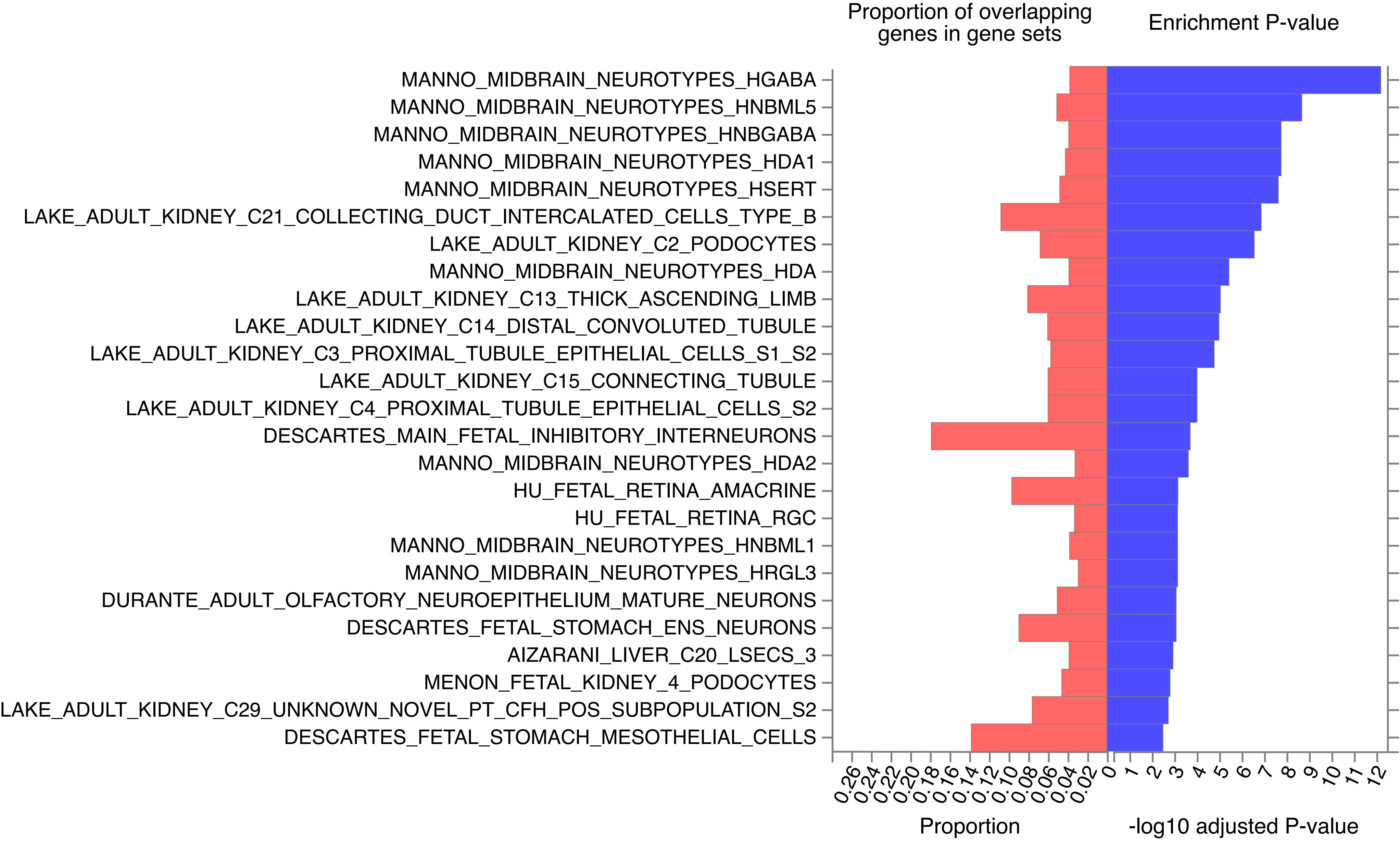

### Figure S3

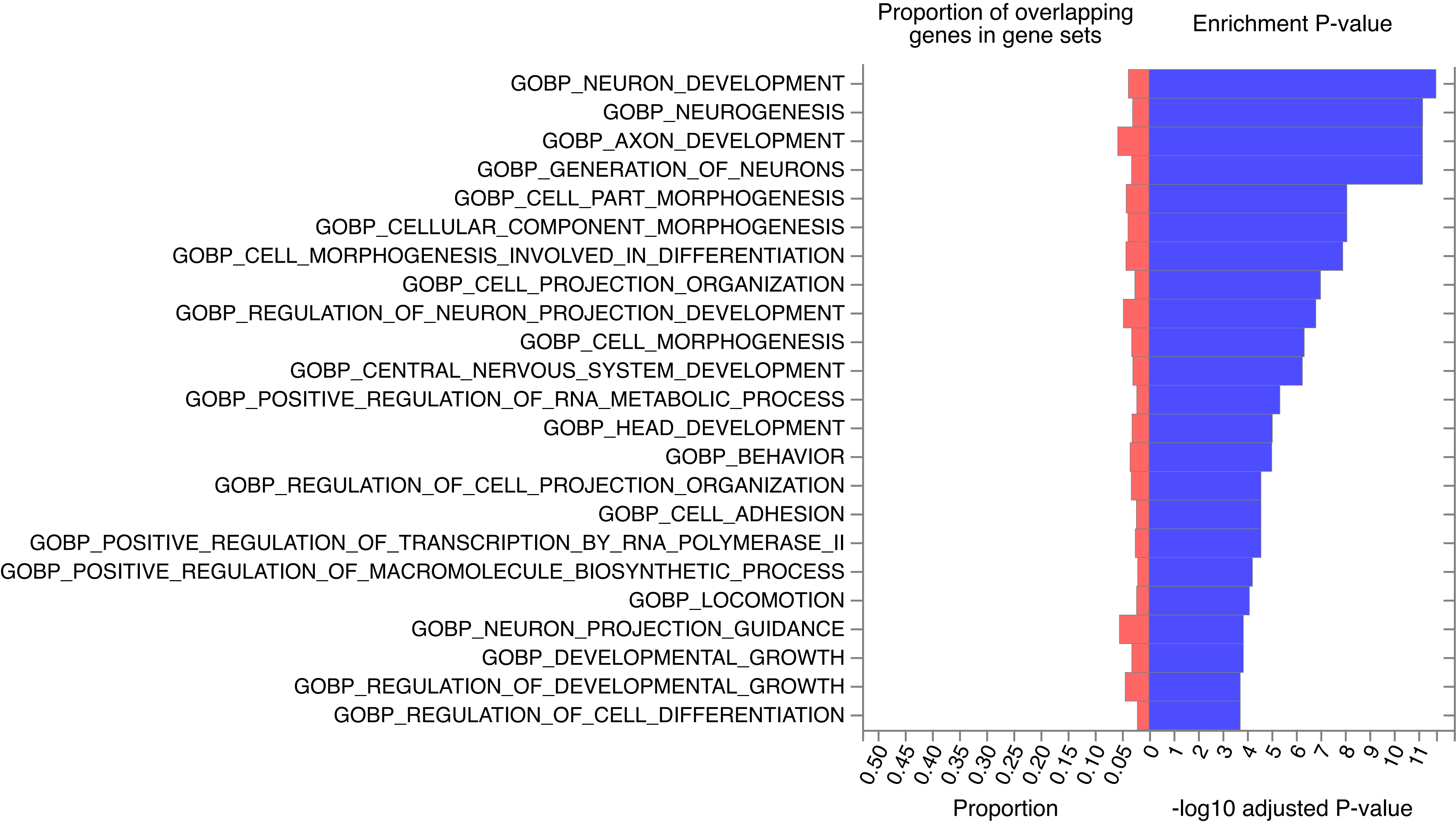

### Figure S4

#### KEGG

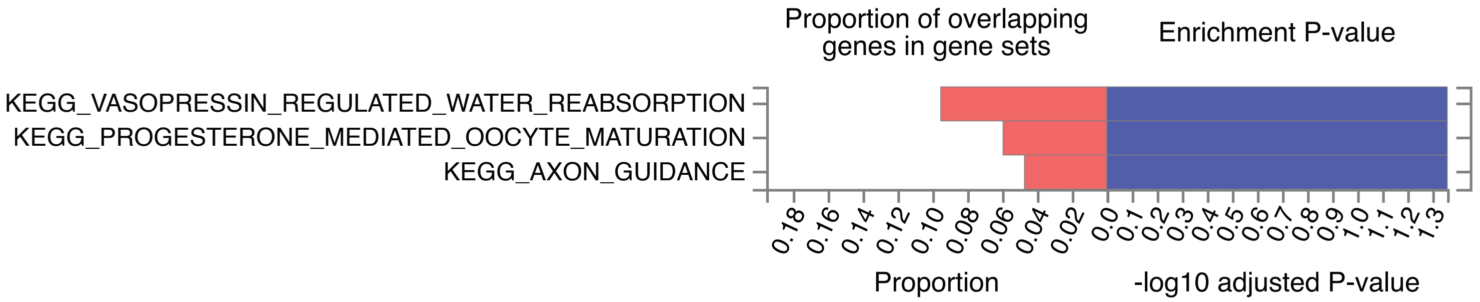


#### Reactome

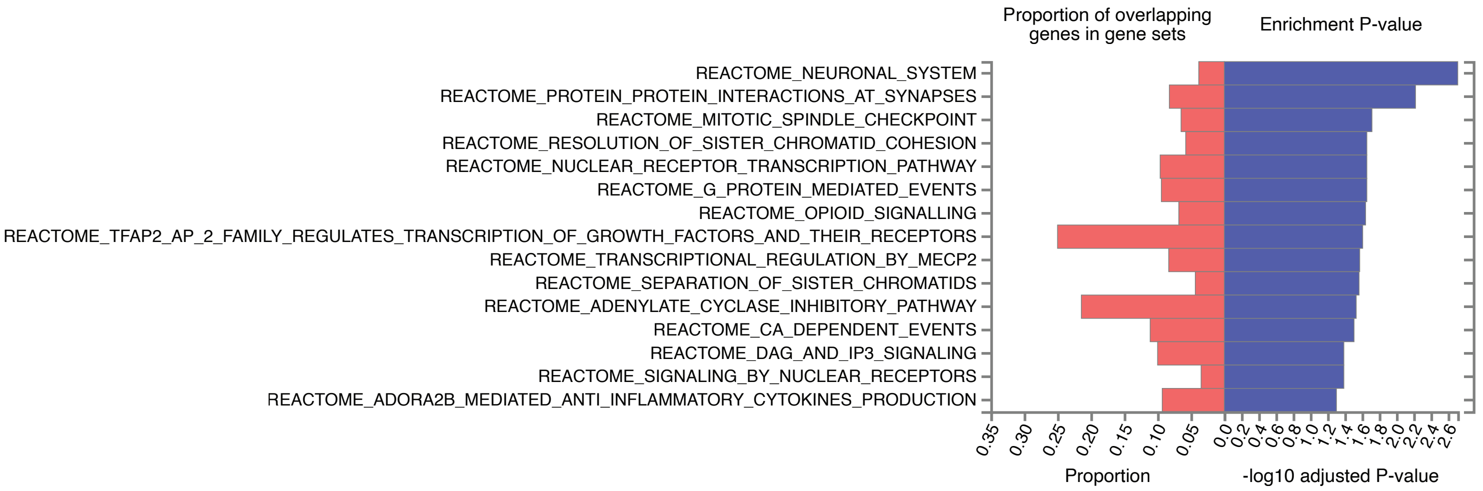


#### WikiPathways

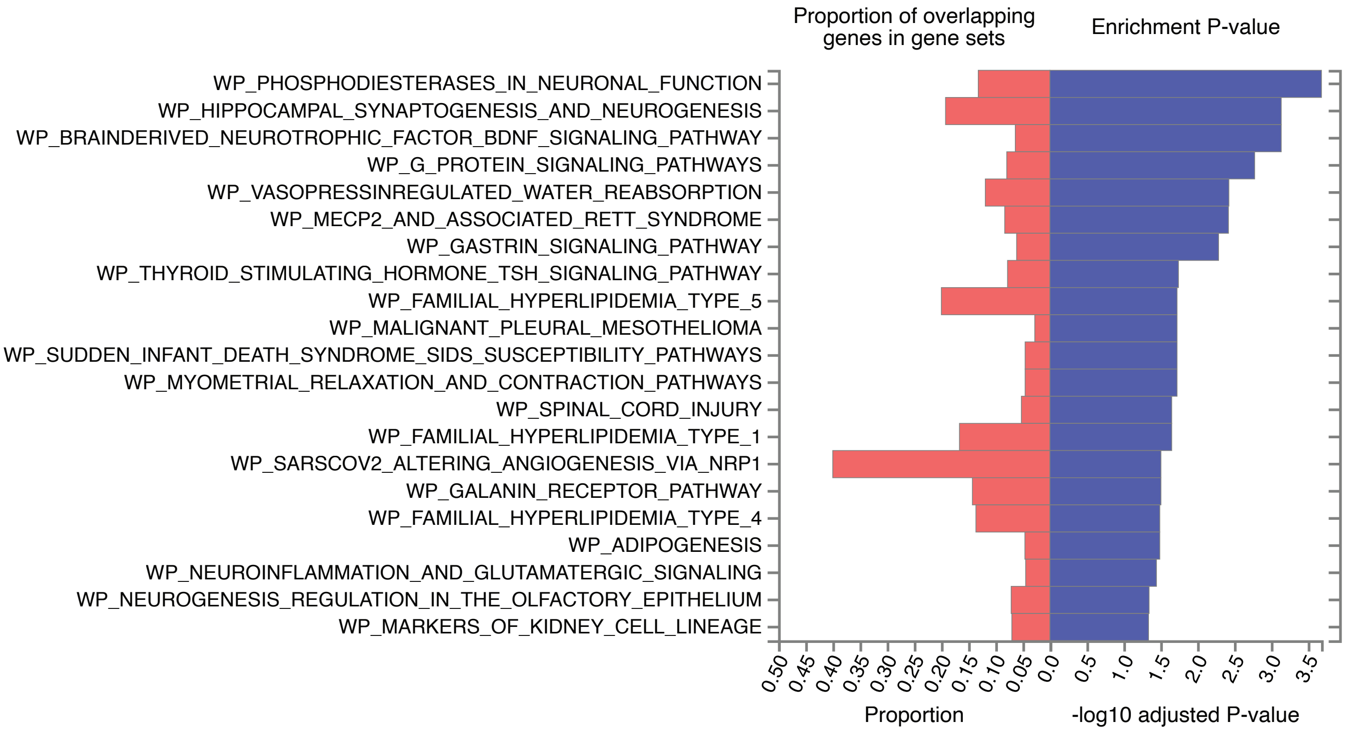

### Figure S5

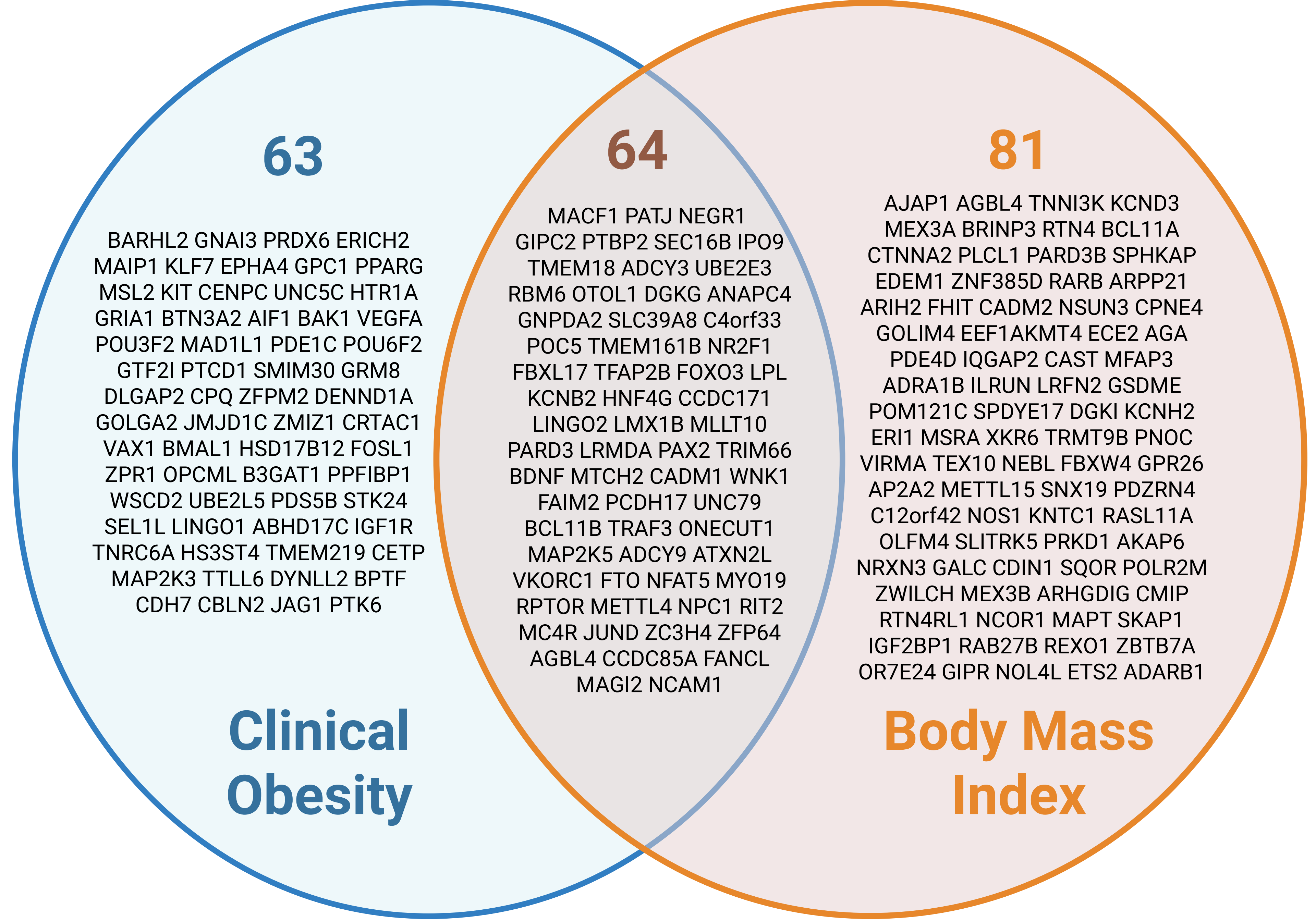

### Figure S6

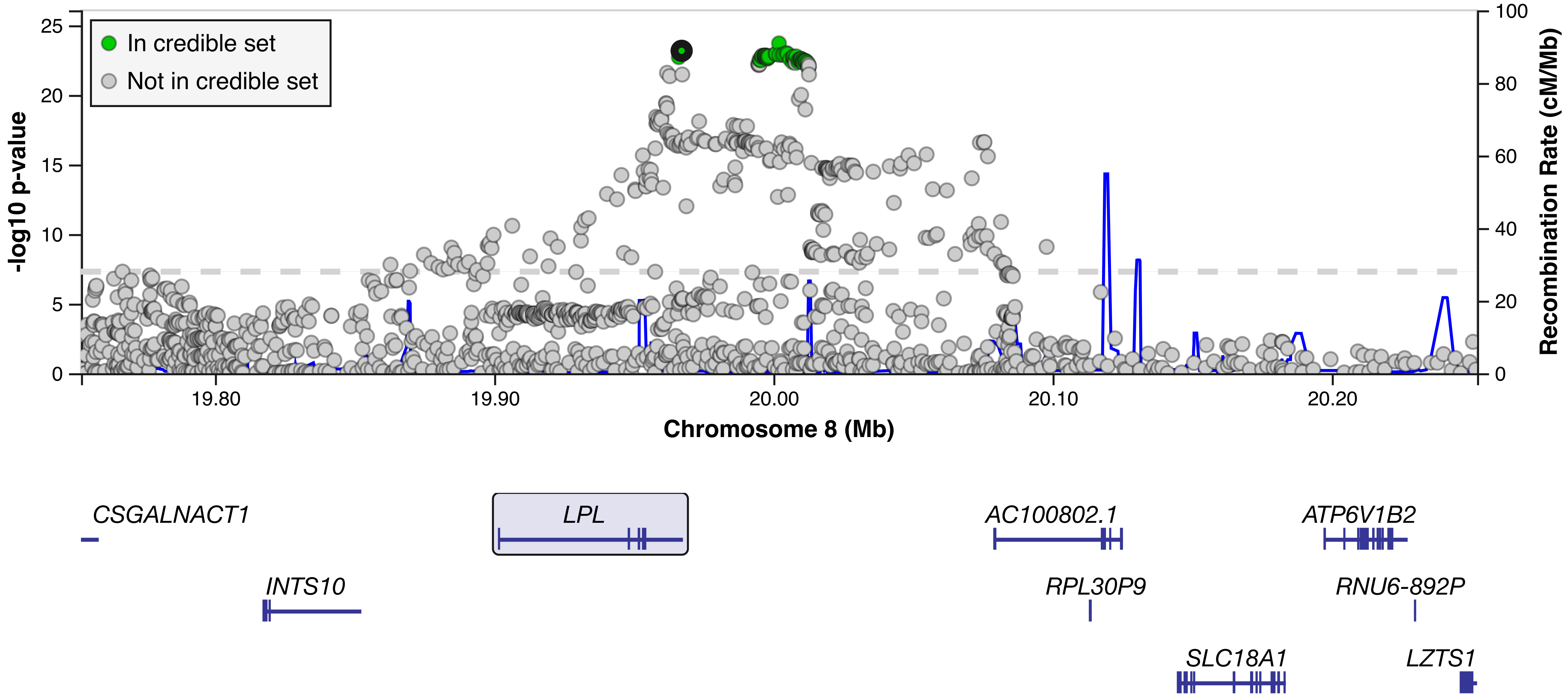

### Figure S7

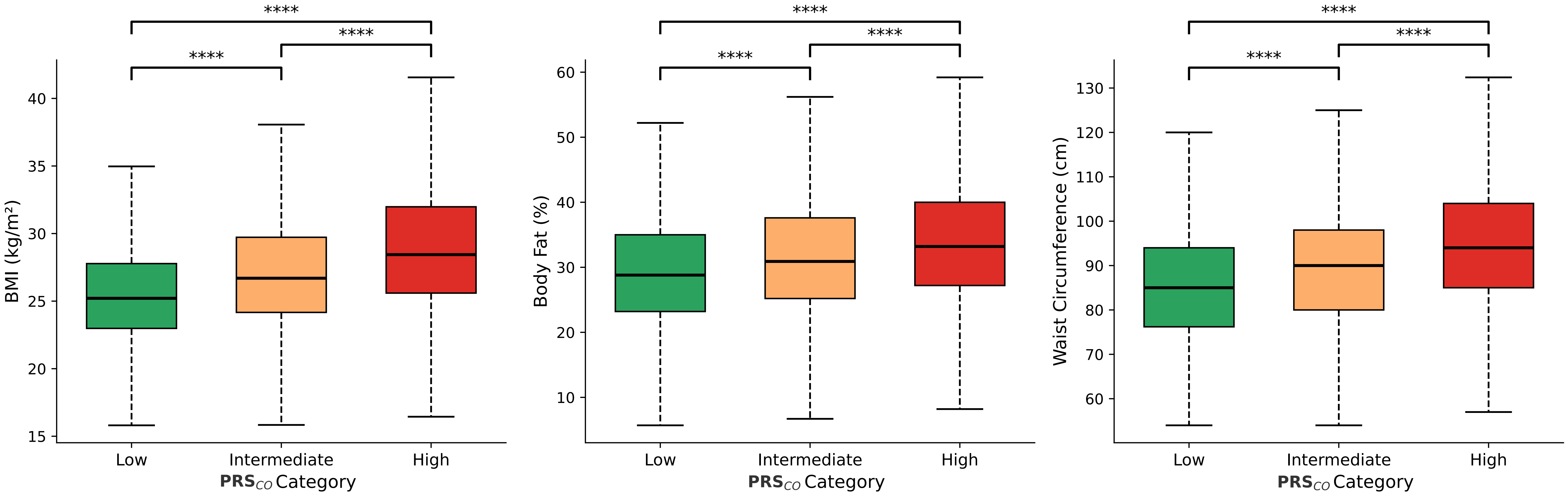

### Figure S8

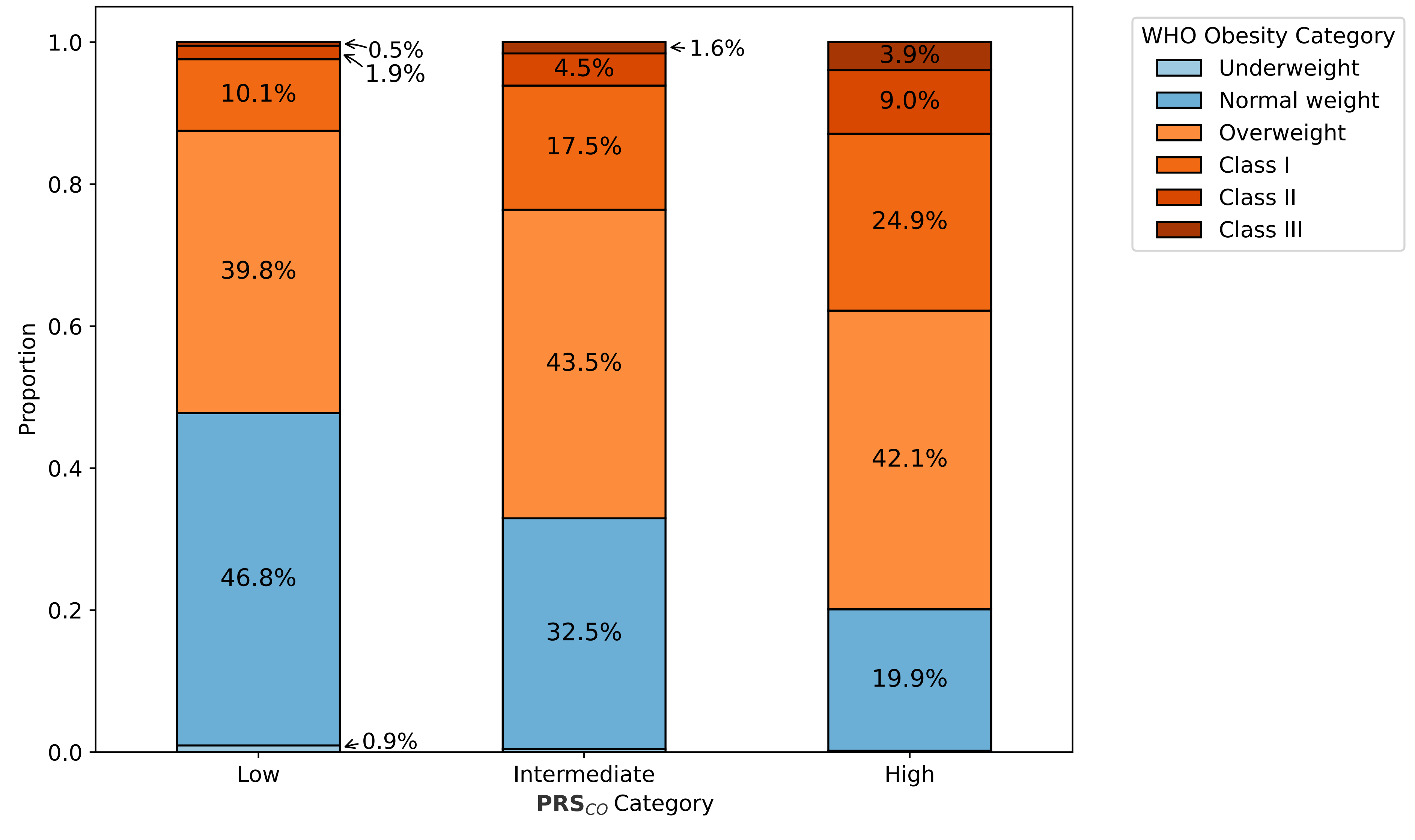

### Figure S9

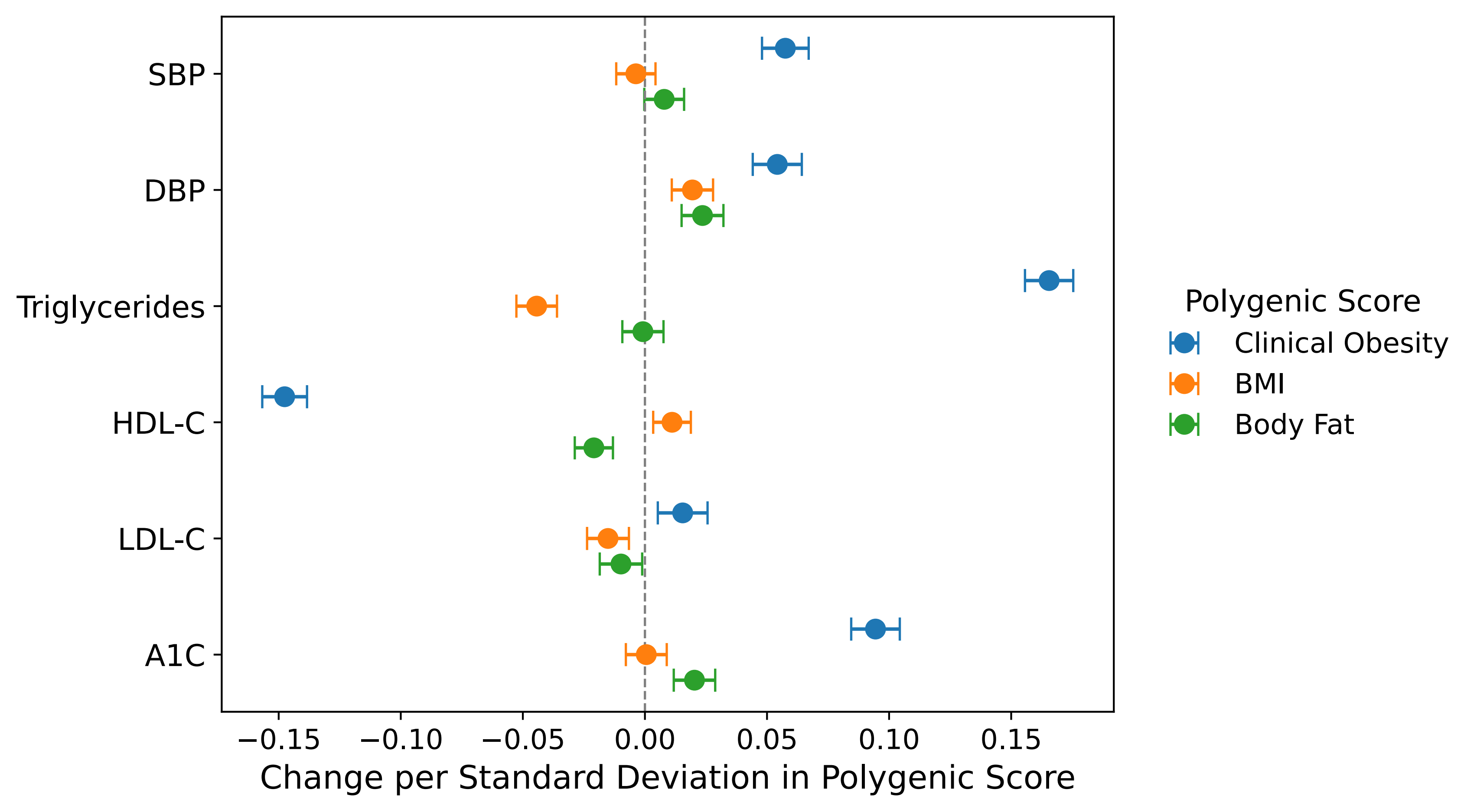

### Figure S10

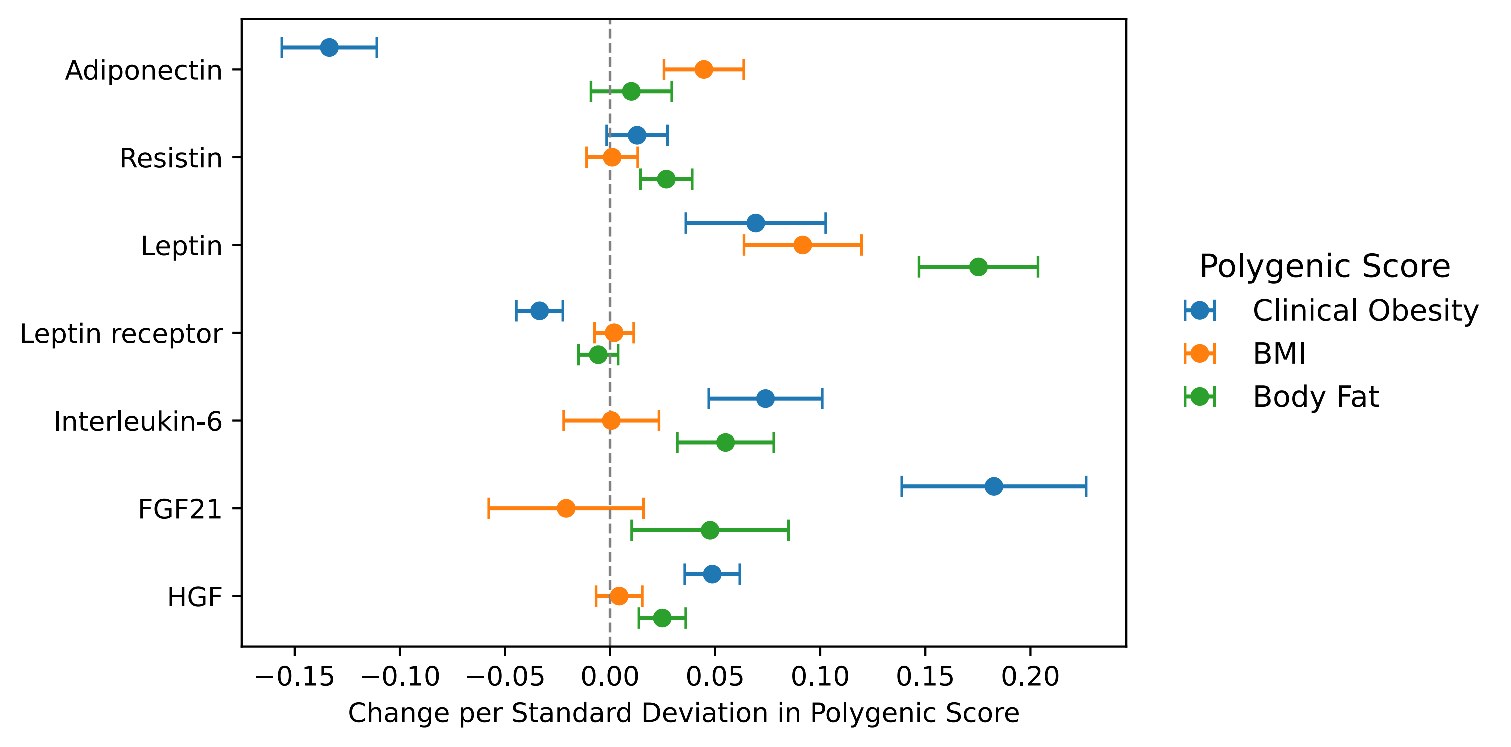

### Figure S11

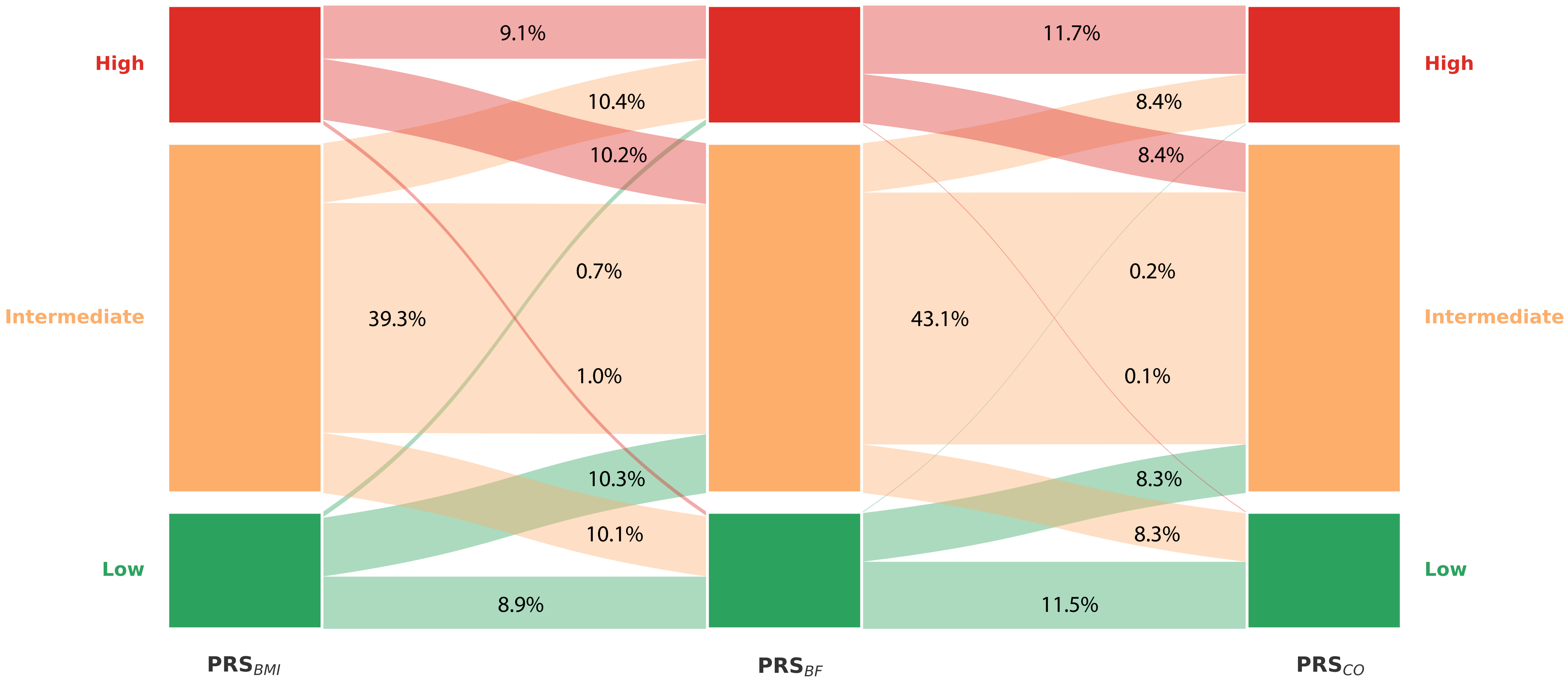

### Figure S12

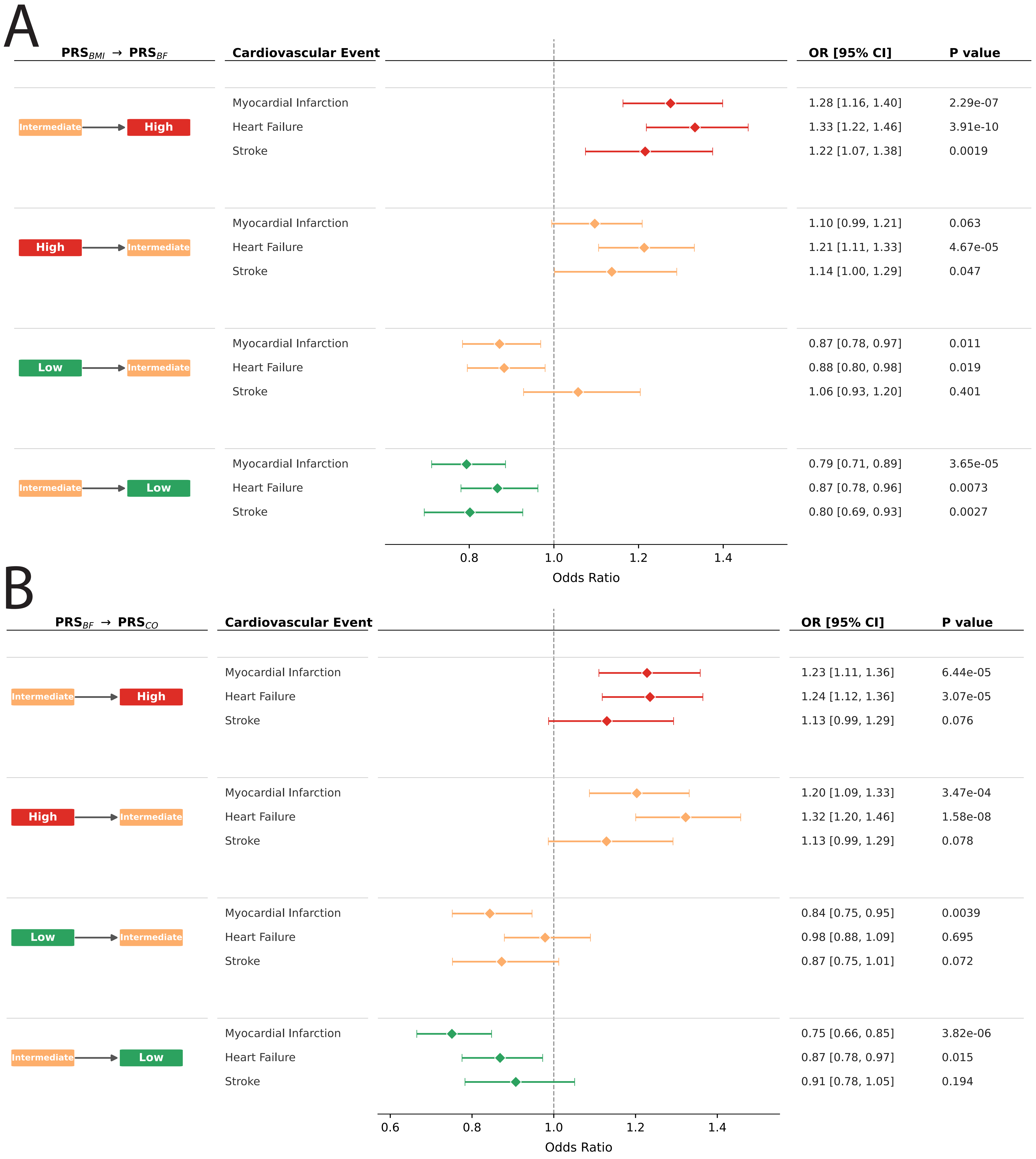

### Figure S13

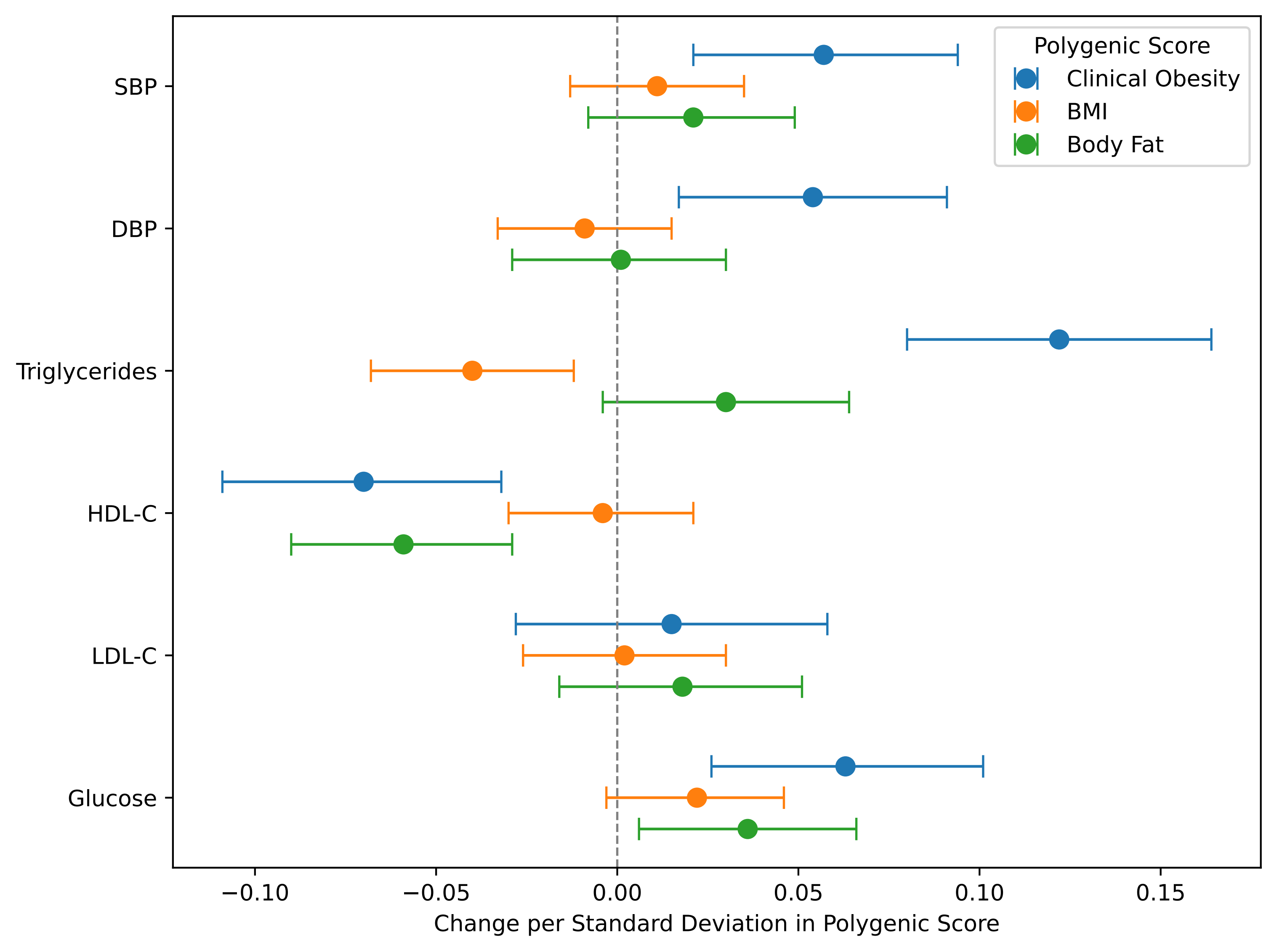
